# Clinical and epidemiological characteristics of children with SARS-CoV-2 infection: case series in Sinaloa

**DOI:** 10.1101/2020.07.07.20146332

**Authors:** Giordano Pérez Gaxiola, Rosalino Flores Rocha, Julio César Valadez Vidarte, Melissa Hernández Alcaraz, Gilberto Herrera Mendoza, Miguel Alejandro Del Real Lugo

**Affiliations:** Department of Evidence-based Medicine. Hospital Pediátrico de Sinaloa “Dr. Rigoberto Aguilar Pico”. Blvd. Constitución y Donato Guerra SN. Culiacán, Sin. C.P. 80200. Culiacán, Sin. México; Department of Epidemiology. Hospital Pediátrico de Sinaloa “Dr. Rigoberto Aguilar Pico”. Blvd. Constitución y Donato Guerra SN. Culiacán, Sin. C.P. 80200. Culiacán, Sin. México; Department of Pediatrics. Hospital Pediátrico de Sinaloa “Dr. Rigoberto Aguilar Pico”. Blvd. Constitución y Donato Guerra SN. Culiacán, Sin. C.P. 80200. Culiacán, Sin. México; Epidemiological Intelligence Unit. Servicios de Salud de Sinaloa, Cerro de Montebello 150, Fracc. Montebello, C.P.80227. Culiacán., Sin. México

**Keywords:** COVID-19, severe acute respiratory syndrome coronavirus 2, epidemiology, Signs and Symptoms

## Abstract

**Background:** The SARS-CoV-2 virus may affect both adults and children. Although the disease, named COVID-19, has a lower prevalence in infancy and has been described as mild, the clinical characteristics may vary and there is a possibility of complications.

**Objectives:** To describe the clinical and epidemiological characteristics of pediatric cases confirmed in the state of Sinaloa, Mexico, during the first three months of the pandemic, and of children admitted with COVID-19 to a secondary hospital.

**Methods:** This case series includes all patients with SARS-CoV-2 infection confirmed by PCR testing, identified in the state epidemiological surveillance system between March 1 and May 31, 2020. Confirmed patients admitted to the Sinaloa Pediatric Hospital (HPS) during the same dates are also described.

**Results:** Fifty one children with SARS-CoV-2 were included, 10 of the admitted to HPS. The median age was 10 years. The more frequent symptoms were fever (78%), cough (67%) and headache (57%). Most cases were mild or asymptomatic. Three patients with comorbidities died. Only 4 of 10 patients identified in HPS had been admitted with the diagnosis of possible COVID-19.

**Conclusions:** SARS-CoV-2 infection in children was mostly mild or asymptomatic, but with a wide range of clinical presentations.

## Introduction

The coronavirus disease (COVID-19) pandemic caused by the SARS-CoV-2 virus, that began in the Wuhan province in China at the end of 2019, has affected more than six million people and has had sanitary and economic repercussions worldwide (1–3). The initial characterization of the disease was described in adults (4) but as the epidemic has advanced neonatal and pediatric cases were identified. (5–8).

Although the prevalence of COVID-19 in childhood represents a low percentage of the totality of reported cases, varying between 0.8% and 2.7% (9–11), the number of children that may become affected and the different clinical presentation of the disease compared to the adult population (4,12) may be a challenge for pediatricians and general practitioners.

Similar to adults, SARS-CoV-2 is more frequent in males. Fever and cough are also the main symptoms but they are less prevalent in childhood. The laboratory and imaging findings appear to be more variable in children. The severity of disease is milder than in adults (9,10,13–15). And, in children, mortality is low and may be lower than that of influenza and other respiratory infections (16). Despite this, the possibility of complications exist (11,17,18), and the possible association of COVID-19 with a severe Kawasaki-like syndrome, named Multisystem Inflammatory Syndrome, is being studied (19–22).

The objectives of this study were to describe the clinical and epidemiological characteristics of pediatric cases confirmed in the state of Sinaloa, Mexico, during the first three months of the pandemic in the region, and of a subset of those children admitted with COVID-19 to Sinaloa Pediatric Hospital (Hospital Pediátrico de Sinaloa “Dr. Rigoberto Aguilar Pico”, HPS).

## Material and methods

During the first three months from the start of the pandemic in Sinaloa, March through May, 2020, clinical and epidemiological data was obtained of infants and children less than 18 years of age with SARS-CoV-2 infection confirmed by polymerase chain reaction testing (PCR) in respiratory secretions. Patients were identified through the System of Epidemiological Surveillance of Respiratory Diseases (SISVER) of the state Health Secretariat. Demographic data, signs and symptoms were recorded at the time of PCR testing. Follow up was attempted with two separate phone calls and a WhatsApp message at least two weeks after the positive test. The same data extraction form was used for those identified that were admitted to HPS. Laboratory and imaging findings, which were ordered at physician’s request, are also described for HPS patients.

Severity of the disease was classified according to the COVID-19 clinical syndromes defined by the World Health Organization (WHO) (23): asymptomatic infection, mild disease (upper respiratory infection), moderate (pneumonia), severe (severe pneumonia), critical (severe acute respiratory syndrome, or sepsis, or septic shock).

### Statistical analysis

Results of clinical and epidemiological characteristics are descriptive, using absolute numbers and percentages, or medians and interquartile ranges as appropriate. Analysis was done using Microsoft Excel version 16.37.

## Results

Between March 1 and May 31, 2020, a total of 3434 SARS-CoV-2 positive cases were confirmed in Sinaloa, 51 (1.5%) of them in children less than 18 years. Of those 51, 26 were males (51%) and the median age was 10 years. In 33 children, a positive close contact was identified.

The most frequent symptoms were fever (78%), cough (67%) and headache or irritability (57%). Less common were odynophagia, rhinorrhea, vomiting and diarrhea. Three patients had cancer, one had chronic kidney disease and hypertension, three had obesity, and one had sequels of head trauma and a lung abscess (Table 1).

**Table 1.**
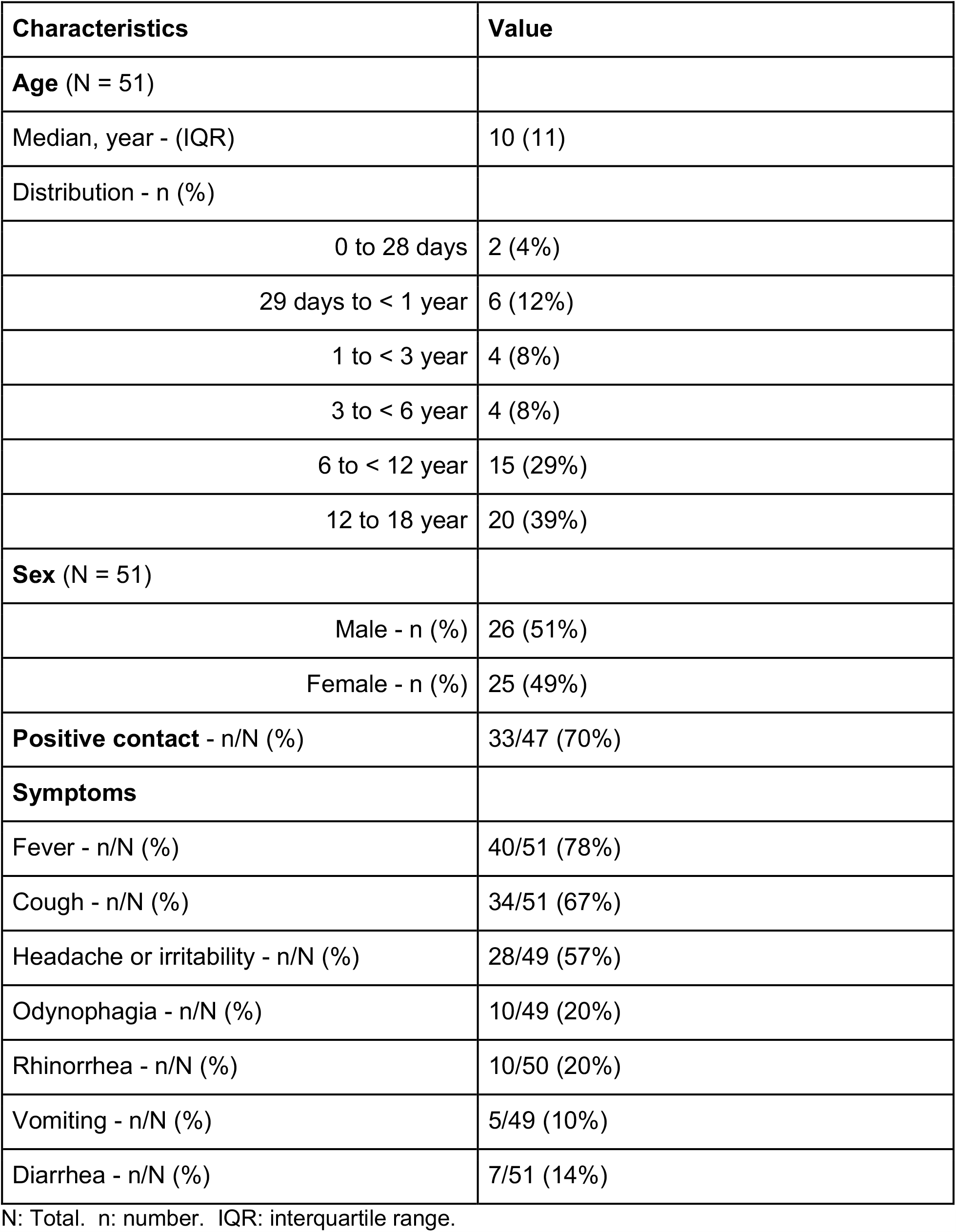
Characteristics of children with SARS-CoV-2 infection in Sinaloa.

The follow up call to record the clinical course and to classify the severity was completed in 37 patients (Table 2). Seven children were asymptomatic(19%), 23 had mild disease (62%), 3 moderate (8%), 1 severe (3%), and 3 critical (8%). There were three deaths, all in children with comorbidities: an infant male with Down syndrome, congenital cardiopathy and hypothyroidism; a school age male with chronic kidney disease and hypertension; and a teen male with sequels of head trauma and a lung abscess.

**Table 2.** Severity of disease in children with SARS-CoV-2 infection in Sinaloa.

| **Category** | **Number (N = 37)** |
| --- | --- |
| Asymptomatic | 7 |
| Mild | 23 |
| Moderate | 3 |
| Severe | 1 |
| Critical | 3 |

Ten of the 51 children were admitted to HPS. Reasons for hospitalization were diverse. In four patients the COVID-19 diagnosis was suspected from the start (patients 5, 6, 8 and 10 in Table 3). The other six had other reasons for admission and developed fever during the hospitalization. One patient with multiple comorbidities died. He had been admitted to the hospital more than two months prior to developing critical COVID-19. The other nine patients had mild or moderate disease, improved and were discharged. Most patients had normal complete blood counts (CBC), with the exception of the three children with neoplastic pathologies. The classical ground glass pattern in lung tomography was observed in six children.

**Table 3.** Characteristics of children with COVID-19 admitted to Hospital Pediátrico de Sinaloa.

| Characteristic | Patient 1 | Patient 2 | Patient 3 | Patient 4 | Patient 5 | Patient 6 | Patient 7 | Patient 8 | Patient 9 | Patient 10 |
| --- | --- | --- | --- | --- | --- | --- | --- | --- | --- | --- |
| Age group | Teenager | Teenager | Infant | School age | Infant | Preschool | Infant | Infant | Preschool | Infant |
| Sex | M | F | M | M | M | M | F | M | F | F |
| Positive contact | No | No | Yes | Yes | Yes | No | Yes | Yes | Yes | Yes |
| Reason for hospitalization | Other | Other | Other | Other | COVID-19 suspicion | COVID-19 suspicion | Other | COVID-19 suspicion | Other | COVID-19 suspicion |
| Comorbidities | Cancer | No | Congenital malformation and genetic syndrome | No | No | Cancer | Congenital malformation | No | All | No |
| Severity | Moderate | Mild | Critical | Mild | Moderate | Mild | Mild | Mild | Mild | Mild |
| Required Oxygen | Yes | No | Yes | No | No | No | No | No | No | No |
| Required ICU | No | No | Sí | No | No | No | No | No | No | No |
| Length of stay (days) | 24 | 21 | 126 | 9 | 8 | 12 | 24 | 6 | 34 | 4 |
| Fever | Yes | Yes | Yes | Yes | Yes | Yes | Yes | Yes | Yes | Yes |
| Duration of fever (days) | 8 | 1 | 2 | 1 | 1 | 1 | 1 | 3 | 3 | 3 |
| Cough | Yes | Yes | Yes | Yes | Yes | Yes | Yes | No | Yes | Yes |
| Cefalea o irritabilidad | Yes | Yes | Yes | Yes | Yes | Yes | Yes | No | Yes | Yes |
| Diarrhea | No | No | No | No | No | Yes | No | No | No | No |
| Abdominal pain | No | No | No | No | No | Yes | No | No | No | No |
| Leucocytes (K/uL) | 240 | 7560 | 13610 | 9470 | 14590 | 80 | 10370 | 11640 | 880 | 31310 |
| Lymphocytes (K/uL) | 230 | 1420 | 1400 | 1988 | 10650 | 0 | 3940 | 1957 | 660 | 10645 |
| C reactive protein (mg/dl) | 29.5 | 10.4 | 1.7 | 0.1 | 0.1 | - | 8 | - | - | 11.6 |
| Procalcitonina (ng/ml) | 0.2 | - | - | - | - | 113.38 | - | 0.08 | 0.29 | - |
| AST (U/L) | 25 | - | 44 | - | - | - | - | - | - | - |
| **ALT (U/L)** | 66 | - | 784 | - | - | - | - | - | - | - |
| **LDH (U/L)** | 131 | - | 470 | - | - | - | - | - | - | - |
| **Chest X-ray** | Ground glass | Perihilar opacity | Ground glass | Normal | Multiple patches | Perihilar opacity | Perihilar opacity | Basal opacity | Perihilar opacity | Perihilar opacity |
| **Lung CT** | Ground glass | Ground glass | - | Ground glass | - | Ground glass | - | Ground glass | Ground glass | - |
| **Clinical course** | Discharged | Discharged | Died | Discharged | Discharged | Discharged | Discharged | Discharged | Discharged | Discharged |

## Discussion

This case series describes the clinical and epidemiological characteristics of children infected by SARS-CoV-2 during the first three months of the pandemic in the state of Sinaloa. The proportion of pediatric cases in relation to the total number was 1.5%, similar to previous reports ranging from 0.8% to 2.7% (9–11).

Also concurring with published case series and systematic reviews, we found more cases in males, more in adolescents and school aged children than in infancy, the severity of disease was mostly mild, and the most frequent symptoms (9,10,13–15). The presence of a ground glass appearance in lung imaging, even in mild or asymptomatic children has also been reported (24–26). No children fulfilling the criteria for Multisystem Inflammatory Syndrome were identified.

The characteristics of the children admitted to HPS highlight the variability that the clinical presentation of COVID-19 may have. Only in four of 10 was COVID-19 suspected at admission. The other six patients were admitted for other reasons, developed fever after hospitalization and then were diagnosed. Other than having a respiratory triage, which HPS implemented from the beginning of the pandemic, the only way to successfully isolate all patients infected from the start would be to screen with PCR testing all children that need hospitalization. This is neither practical nor economically feasible because the prevalence of COVID-19 is much lower in children than in adults (27), and because of the availability and high cost of tests. The identification of seven asymptomatic children state wide also may have implications of the transmissibility in the community and should be considered for epidemiological surveillance.

This study has weaknesses. First, we could only contact 70% of patients in the two week follow up despite calling by phone two times and sending messages using WhatsApp. Second, the reporting of symptoms at the moment of diagnosis was subjective, although since it is an acute disease recall bias is less likely. Third, data from laboratory and imaging tests was limited to patients in HPS and were ordered by attending physicians. The findings could have varied in children that were not hospitalized or in the few that were admitted to other institutions.

Although the majority of patients in this series had no symptoms, or mild or moderate disease, the possibility of complications exist, mainly in children with comorbidities (11,17,18). The three patients that died had concomitant pathologies. The proportion of severe or critical disease should remain low in the next months of the pandemic but considering that cases are still increasing this may represent a logistical challenge for pediatric and general hospitals. The two positive neonates also highlight that, although infrequent, neonatal infection may develop and should be considered in hospital with maternal wards. Fortunately, though, the burden of COVID-19 in pediatric wards should be less than what the influenza virus annually causes (16).

## Data Availability

Available per request.

## Acknowledgments

We thank Dr. Rafael Félix Espinoza and M.S.P. Silvia Karina Meza Rodríguez, for their support.

